# Delivery mode impacts gut bacteriophage colonization during infancy

**DOI:** 10.1101/2023.11.13.23298307

**Authors:** Poorani Subramanian, Hector N. Romero-Soto, David B. Stern, George L. Maxwell, Shira Levy, Suchitra K. Hourigan

## Abstract

**Background:** Cesarean section delivery is associated with altered early-life bacterial colonization and later adverse inflammatory and immune health outcomes. Although gut bacteriophages can alter gut microbiome composition and impact host immune responses, little is known about how delivery mode impacts bacteriophage colonization over time. To begin to address this we examined how delivery mode affected bacteriophage colonization over the first two years of life.

**Results:** Shotgun metagenomic sequencing was conducted on 272 serial stool samples from 55 infants, collected at 1-2 days of life and 2, 6, 12 and 24 months. 33/55 (60%) infants were born by vaginal delivery. DNA viruses were identified, and by host inference, 94% of the viral sequences were found to be bacteriophages. Alpha diversity of the virome was increased in vaginally delivered infants compared to cesarean section delivered infants at 2 months (Shannon index, p=0.022). Beta diversity significantly differed by delivery mode at 2, 6, and 12 months when stratified by peripartum antibiotic use (Bray–Curtis dissimilarity, all p<0.05). Significant differentially abundant predicted bacteriophage hosts by delivery mode were seen at all time points. Moreover, there were differences in predicted bacteriophage functional gene abundances up to 24 months by delivery mode. Many of the functions considered to play a role in host response were increased in vaginal delivery.

**Conclusions:** Clear differences in bacteriophage composition and function were seen by delivery mode over the first two years of life. Given that phages are known to affect host immune response, our results suggest that future investigation into how delivery mode may lead to adverse inflammatory outcomes should not only include bacterial microbial colonization but also the potential role of bacteriophages and transkingdom interactions.

## BACKGROUND

Infancy represents a key window for gut microbiome establishment and plays a critical role in immune and metabolic education [1-3]. Disturbing the timing and order of microbial colonization can have lasting immune and inflammatory consequences [4, 5]. Cesarean section (CS) delivery represents an important factor perturbing the bacterial colonization of an infant and has been associated with later risk of several inflammatory conditions [6-8]. We read with interest the recent work of Shah *et al*, revealing previously unappreciated virome diversity in the infant gut microbiome at 1 year of age, and uncovering unknown viral species and virus family-level clades [9]. Although gut bacteriophages can alter microbiome composition and function and directly affect host immune responses [10-13], the effect of delivery mode has largely been limited to investigation of bacterial taxa. Transmission of the gut virome from mother to infant has shown lower transmission of viral communities than bacterial communities, and that virome colonization was determined more by dietary and environmental factors rather than direct maternal acquisition [14, 15]. In the first week of life, delivery mode did not determine how much of the virome was shared between mother and infant but some effect of delivery mode on bacteria-bacteriophage interaction was seen [14]. However, how delivery mode affects virome colonization longitudinally in infancy and early childhood has not been studied. Therefore, building on this previous work, we aimed to assess the impact of delivery mode on the infant gut virome over the first two years of life.

## METHODS

### Subjects, sample, and clinical data collection

Mothers were enrolled prenatally with informed consent in an Institutional Review Board approved longitudinal, prospective cohort study “The First 1000 Days of Life and Beyond” (Inova protocol #15-1804, WCG protocol #20120204). Serial stool samples were collected from infants at 1-2 days after birth and at around 2, 6, 12, and 24 months of age as previously described [16, 17]. All samples except for the 1-2 day samples were collected at home and mailed back to the lab, using previously validated methods and stored at -80°C until analysis [18]. Subjects for this current study were chosen from the larger cohort if stool samples were available at all time points. Demographic information, pregnancy details including mode of delivery mode and maternal antibiotic use, and infant data including feeding mode and antibiotic use were collected through a questionnaire, review of electronic medical records and serial surveys. Correlation between clinical and demographic factors with delivery mode was evaluated using the Chi-square test and two-sample t-test for categorical and continues variables respectively.

### DNA extraction and Shotgun metagenomic sequencing

DNA was extracted from stool aliquots using the DNeasy PowerSoil Pro kit (Qiagen, Valencia, CA) following manufacturer’s instructions. Shotgun metagenomic sequencing was performed on the Novaseq platform (Illumina, CA, USA). Positive controls (DNA sequences) and negative controls (DNA free water) were used.

### Virome and bacteriome analysis

Read pairs were trimmed with BBDuk v38.0.1 and then assembled using metaSPAdes. Contigs were then processed following a standardized protocol using VirSorter, CheckV, and DRAM-v to identify viral sequences [19]. These sequences were then clustered using VContact. Bacterial hosts of phage sequences were identified using Phist. Putative genes identified by DRAM-v were annotated by aligning to NCBI’s nr database using DIAMOND [20]. DRAM-v was also used to identify viral auxiliary metabolic genes (vAMGs). Taxonomic classification of the whole microbiota, including the bacteriome was done using Kraken v2. Beta diversity was compared between groups using PERMANOVA with the adonis function from the vegan R package. All other statistics were done with linear mixed effects models using the lmerTest R package on its own and through the MaAsLin2 R package with log transformation to assess differential abundances of virome features.

## RESULTS AND DISCUSSION

Shotgun metagenomic sequencing was conducted on 272 serial stool samples from 55 infants, collected at 1-2 days of life and 2,6,12, and 24 months. 33/55 (60%) infants were born by vaginal delivery (VD). CS delivered infants were more likely to have exposure to maternal peripartum antibiotics (p<0.001). There were no differences by delivery mode in sex, ethnicity, race, breast feeding and infant antibiotic use (Table S1).

From the virome analysis of the sequence data, we identified DNA viruses and attempted to resolve taxonomies. 9745 unique viral clusters were identified. Of these, only 2.2% could be identified, which were mainly bacteriophages. Clusters from the Siphoviridae family were the most common (64%), followed by Myoviridae (28%) and Podoviridae (8%). After aligning the putative gene sequences to NCBI’s nr database, we had an increased classification of 94% of putative gene sequences on average for each time point. The number of putative viral genes increased over time (p=5.05e-71, Fig. S1).

We then assessed the alpha and beta diversity of the virome by delivery mode and time. Alpha diversity (Shannon index) increased over time (p=0.022, Fig. 1A). This differs from some previous studies which report decreasing bacteriophage diversity over the first few years of life, possibly due to differences in preparation of the samples and updated bioinformatics tools and databases utilized in our study [15, 21]. However other reports concur with our study and show an increasing diversity of bacteriophages in infancy [22, 23]. There was increased alpha diversity in VD compared with CS at birth (p= 0.0028) and 2 months (p=0.009), with significance remaining after adjusting for peripartum antibiotic use. No difference in alpha diversity by delivery mode was seen at later timepoints. Interestingly, the only other study examining the virome by delivery mode later in infancy in 20 infants at a single time point of 12 months, still found increased alpha diversity in those vaginally delivered at 1 year [24]. In our study the analysis at birth is somewhat limited as fewer CS samples could be used (n=6) due to poor quality reads and/or no identifiable viral sequences. Mother-infant vertical transmission of the virome is thought to be lower than the bacteriome [14]; our results suggest this may be further lowered by CS. No differences were seen in alpha diversity by other clinical factors including breast feeding and infant antibiotic use, although others have reported that breast feeding can alter infant virome colonization [23]. Beta diversity (Bray-Curtis) differed by timepoint (p< 0.005, Fig. 1B) but not overall by delivery mode. However, within each time point, beta diversity was significantly different by delivery mode when stratified by peripartum antibiotic use at 2 (p=0.038), 6 (p=0.002), and 12 months (p=0.039).

**Figure 1:**
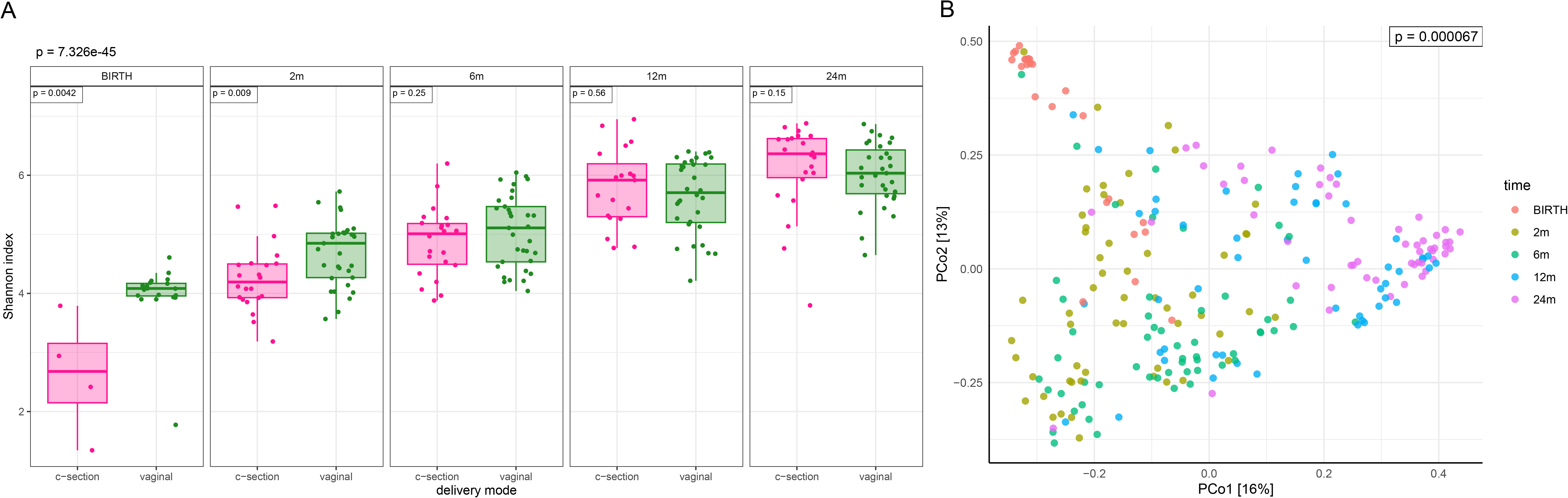
**A:** Alpha diversity (Shannon index) of the virome over time and by delivery mode. **B:** Beta diversity (Bray–Curtis dissimilarity) of the virome over time.

We also assessed the phage abundances and putative host bacterial taxa abundance over time and delivery mode. The vast majority of viral clusters were from bacteriophages (94%). 10 identified phages were shown to significantly differ by delivery mode (FDR < 0.1) at 2 months (Fig. 2A), but not at later timepoints; all were higher in VD than CS. Certain predicted host bacterial species differed significantly in abundance by delivery mode at each time point after birth, with an increased number of significantly differentially abundant hosts identified in VD compared with CS up to 12 months (Fig. 2B). Notably, phages belonging to the *Bacteroidaceae* hosts were increased in VD at 2 and 6 months, with the bacterial taxa *Bacteroides* known to be increased in VD compared to CS infants in early life [25]. At 24 months, there were more differentially abundant phage hosts predicted in CS compared to VD. The predicted host bacterial taxa correlated closely with the bacterial abundance of the whole microbiome at all timepoints (Fig. S2).

**Figure 2:**
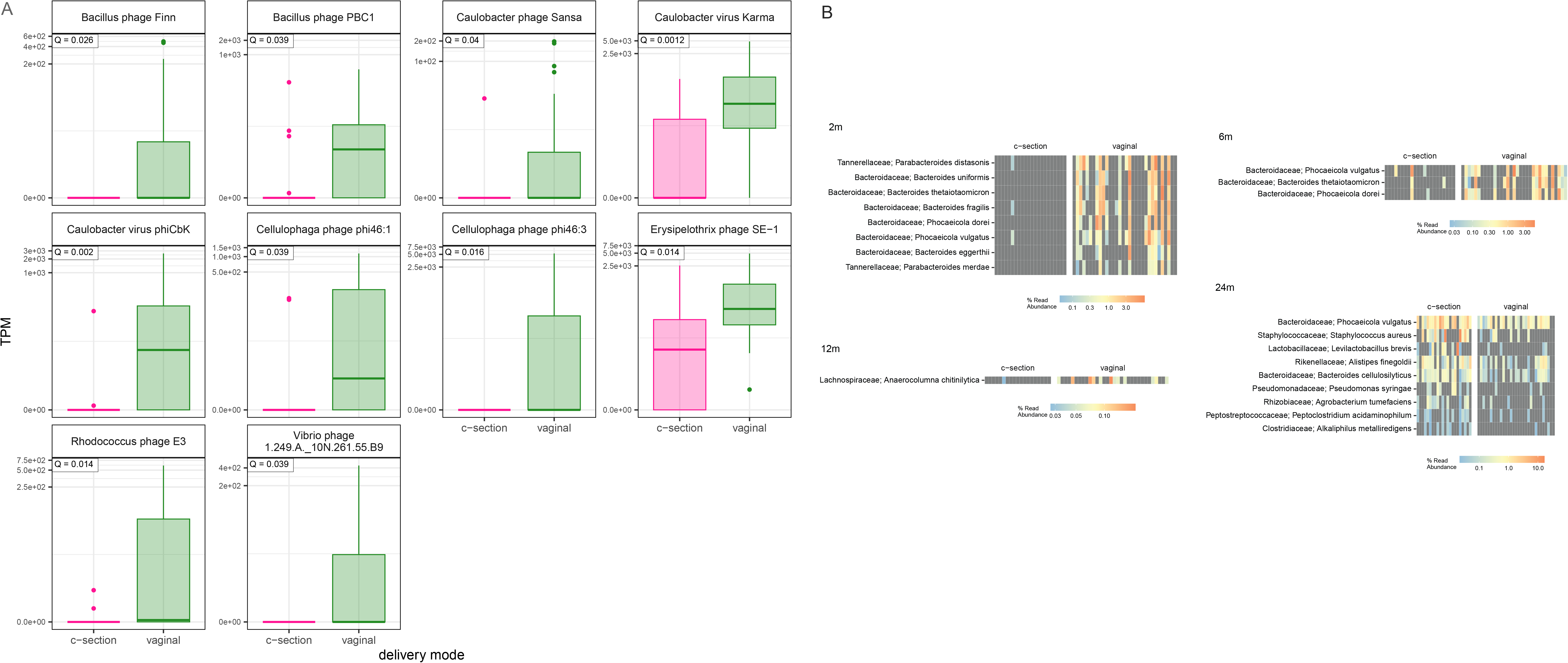
**A**: Significantly differentially abundant bacteriophages by delivery mode at 2 months. **B:** Significantly differentially abundant predicted bacterial hosts over time and by delivery mode.

We then attempted to assess the predicted functional genes and pathways of the identified virome over time and by delivery mode. Very few functional pathways were identified and did not differ by delivery mode. However, when looking at vAMGs, differences were found by delivery mode at birth, 2, 6, and 24 months. Of the differentially abundant gene sequences, only fabG, associated with sugar metabolism in pathogenic bacteria, was found to be in higher abundance in CS than VD at 2m [26]. All others were higher in abundance in VD compared to CS and included ENO, tkA, Glycos_transf_2, PGD, UGDH, ABC.CD.A and RPL19, genes associated with human host response. Others of note, essential to bacteria, included SusC, iroN, DNMT1 and UGP2. Full results are shown in Figure S3 and Table S2.

The reasons for the difference in bacteriophage colonization and function by delivery mode up to 24 months is still yet to be determined. While differential acquisition of the maternal virome is possible, as infants delivered by CS bypass the birth canal and maternal vaginal and perineal microbiome exposure, studies suggest that maternal to infant transmission of the virome in early life is already very low [14, 15]. It has been suggested dietary factors such as breastfeeding contribute to early life virome colonization, however there were no differences in breastfeeding by delivery mode in our study (Table S1). It is likely that the differential bacterial colonization known to occur by delivery mode also affects bacteriophage differences, as evidenced by predicted bacteriophage hosts closely resembling whole microbiome bacterial composition (Fig. S2) [6, 27-29]. In addition, peripartum antibiotic exposure, which is higher in CS delivery, may play a role in bacteriophage colonization as suggested by a difference in virome composition (beta diversity) by delivery mode when stratified by peripartum antibiotic use.

Although to our knowledge, our study is the first to examine the impact of delivery mode on infant virome colonization over the first 2 years of life, there are several limitations. Only DNA viruses were examined and so how delivery mode may impact gut RNA viruses remains unknown. Additionally, maternal vaginal and stool virome were not assessed and so maternal to infant transmission by delivery mode cannot be assessed.

## CONCLUSIONS

Clear differences in bacteriophage composition and function were seen by delivery mode over the first two years of life. Given that phages are known to affect host immune response, our results suggest that future investigation into how delivery mode may lead to adverse inflammatory outcomes should not only include bacterial microbial colonization but also the potential role of bacteriophages and transkingdom interactions.

## Data Availability

The data sets generated and/or analyzed in the current study are available in the NCBI SRA repository (https://dataview.ncbi.nlm.nih.gov/object/PRJNA988496?reviewer=usujv2ng73h0ftdbiavm8soe98) under BioProject number: PRJNA988496.

## ABBREVIATIONS

CS: Cesarean section
VD: Vaginal delivery
FDR: False discovery rate
vAMGs: Viral auxiliary metabolic genes
fabG: 3-oxoacyl-[acyl-carrier-protein] reductase FabG
ENO: Enolase
tkA: Transketolase
Glycos_transf_2: Glycosyl transferase family 2
PGD: 6-phosphogluconate dehydrogenase
UGDH: UDP-glucose 6-dehydrogenase
ABC.CD.A: Putative ABC transport system ATP-binding protein
RPL19: Large subunit ribosomal protein L19
SusC: TonB-dependent starch-binding outer membrane protein SusC
iroN: Iron complex outermembrane recepter protein
DNMT1: DNA (cytosine-5-)-methyltransferase
UGP2: UTP--glucose-1-phosphate uridylyltransferase

## ACKNOWLEDGEMENTS

Inova expresses its appreciation to the Fairfax County in VA, which has supported Inova’s research projects with annual funding from its Contributory Fund (Fund 10030, Maxwell). This work also used the Office of Cyber Infrastructure and Computational Biology High Performance Computing cluster at NIAID, Bethesda, MD.

## AUTHOR INFORMATION

Poorani Subramanian and Hector N. Romero-Soto contributed equally to this paper.

## Authors and Affiliations

National Institute of Allergy and Infectious Diseases, National Institutes of Health, Bethesda, Maryland, USA

Poorani Subramanian, Hector N. Romero-Soto, David B. Stern, Shira Levy & Suchitra K. Hourigan

Women’s Service Line, Inova Health System, Falls Church, Virginia, USA

George L. Maxwell

## Contributions

S.K.H., G.L.M. and S.L. conceived and designed the project. All authors acquired, analyzed, or interpreted data from the study. P.S., H.N.R.-S. and D.B.S. performed statistical analysis. S.K.H. and G.L.M. obtained funding for the project. S.K.H. supervised the project. P.S., H.N.R.-S. and S.K.H. wrote the original draft. All authors reviewed the manuscript for important intellectual content and approved the final manuscript.

## ETHICS DECLARATIONS

### Ethics approval and consent to participate

The study was approved by an Institutional Review Board (Inova protocol #15-1804, WCG protocol #20120204) and all participants provided written informed consent to participate.

### Consent for publication

Not applicable.

## SUPPLEMENTARY INFORMATION

**Table S1:** Demographic and clinical factors stratified by delivery mode.

**Table S2:** Functions of differentially abundant viral auxiliary metabolic genes (vAMGs) by delivery mode

**Figure S1:** Identified viral genes over time.

**Figure S2**: Predicted host bacterial abundance and whole microbiome bacterial abundance at a phylum level

**Figure S3A-D:** Differentially abundant viral auxiliary metabolic genes (vAMGs) by delivery mode and timepoint.

