## Supplementary figures and images for "Delivery mode impacts gut bacteriophage colonization during infancy"

### Supplemental Figure 1

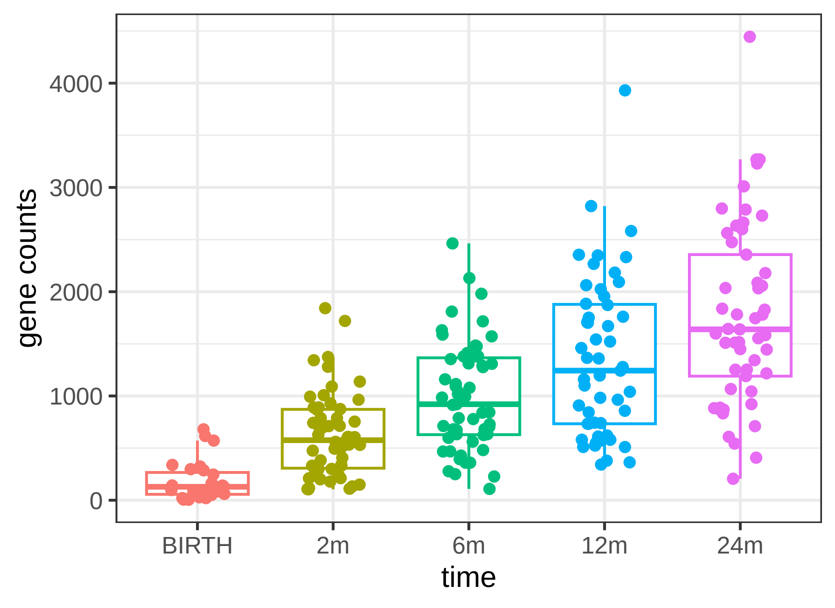

### Supplemental Figure 2

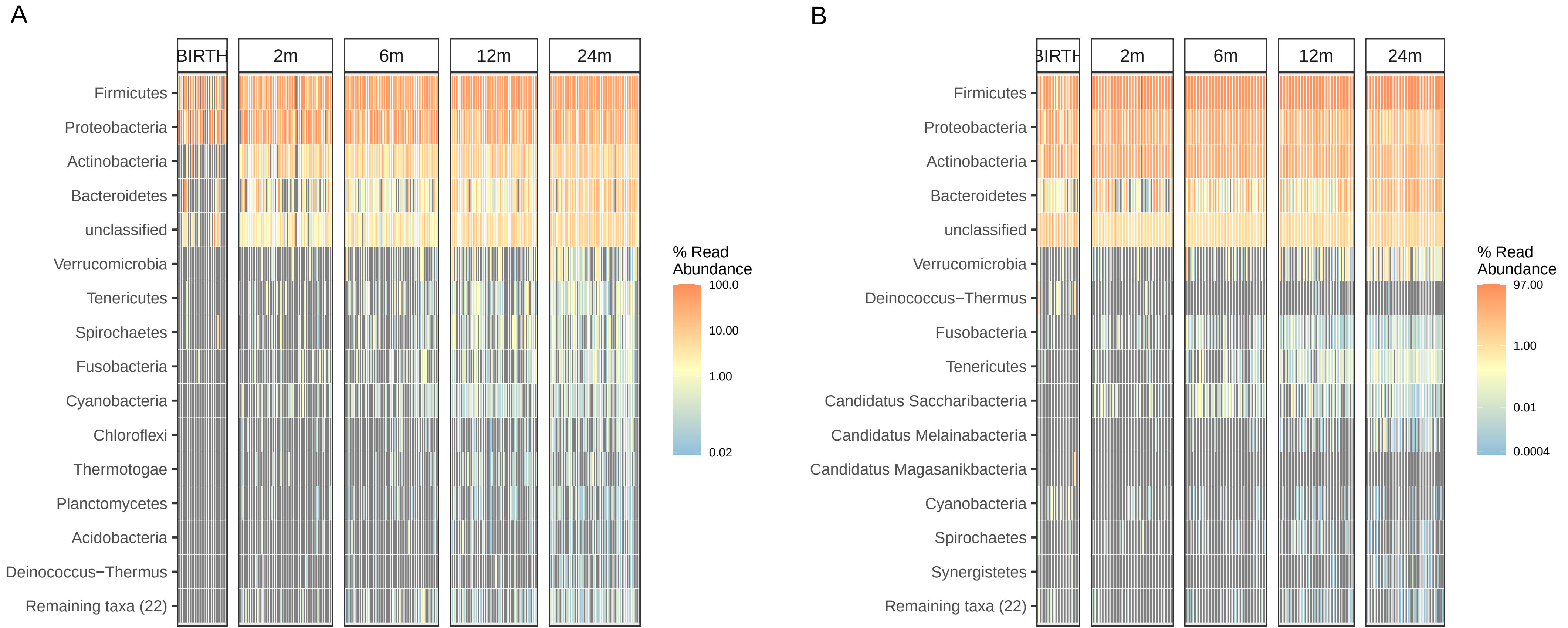

### Supplemental Figure 3

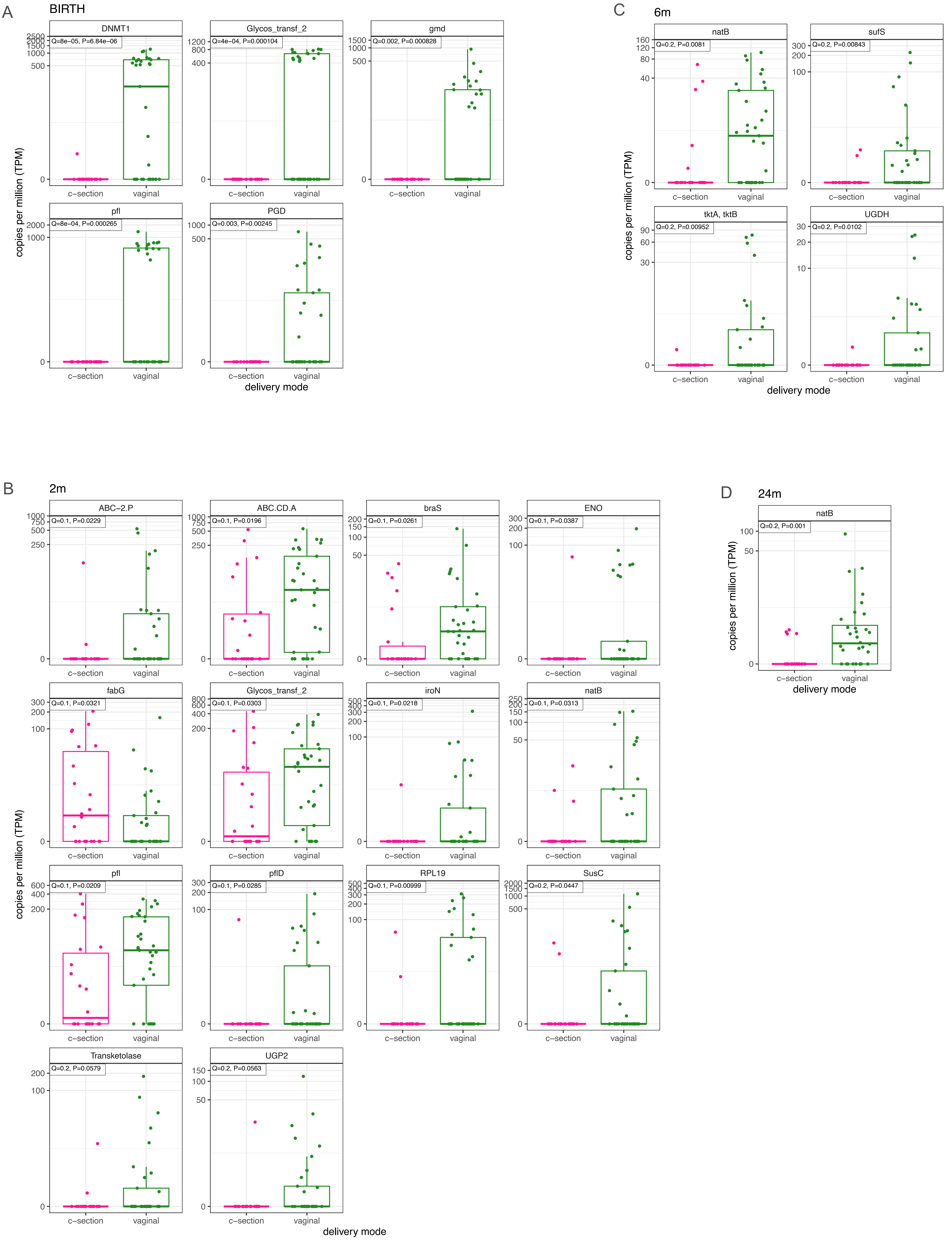
