## Supplemental Table 1 for "Delivery mode impacts gut bacteriophage colonization during infancy"

| Clinical factor | Vaginal delivery (n=33) | Cesarean Section (n=22) | P value |
|---|---|---|---|
| **Sex (n, %)** | | | |
| Male | 18, 55% | 10, 45% | 0.509 |
| Female | 15, 45% | 12, 55% | |
| **Any Breast feeding at 6 months (n, %)** | | | |
| Yes | 25, 76% | 15, 68% | 0.742 |
| No | 5, 15% | 4, 18% | |
| Unknown | 3, 9% | 3, 14% | |
| **Number of breast feeds per week at 6 months** | | | |
| Mean | 29.87 | 31.68 | 0.167 |
| Range | 0 – 99 | 0 – 70 | |
| **Ethnicity (n, %)** | | | |
| Not Hispanic or Latino | 31, 94% | 21, 95% | 1 |
| Hispanic or Latino | 2, 6% | 1, 5% | |
| **Race (n, %)** | | | |
| White or Caucasian | 21, 64% | 17, 77% | 0.197 |
| Black or African American | 5, 15% | 1, 4.5% | |
| Asian | 6, 18% | 1, 4.5% | |
| More than one Race | 1, 3% | 2, 9% | |
| Other | 0, 0% | 1, 4.5% | |
| **Maternal peripartum antibiotics (n, %)** | | | |
| Yes | 7, 21% | 22, 100% | **<0.001** |
| No | 26, 79% | 0, 0% | |
| **Infant antibiotics prior to 2 months (n, %)** | | | |
| Yes | 3, 9% | 4, 18% | 0.419 |
| No | 30, 91% | 18, 82% | |
| **Infant antibiotics prior to 6 months (n, %)** | | | |
| Yes | 4, 12% | 6, 27% | 0.175 |
| No | 29, 88 % | 16, 73% | |
| **Infant antibiotics prior to 12 months (n, %)** | | | |
| Yes | 13, 39% | 10, 45% | 0.655 |
| No | 20, 61% | 12, 55% | |
| **Infant antibiotics prior to 24 months (n, %)** | | | |
| Yes | 15, 45% | 11, 50% | 0.741 |
| No | 18, 55% | 11, 50 % | |
