## Supplemental Table 1 for "Delivery mode impacts gut bacteriophage colonization during infancy"

| Gene | Symbol | Relevant Characteristic(s) | Increased in VD | References |
|---|---|---|---|---|
| 3-oxoacyl-[acyl-carrier-protein] reductase FabG | fabG | Essential for survival in *Escherichia coli* | No (2 m) | 1 |
| TonB-dependent starch-binding outer membrane protein SusC | SusC | Essential for utilization of maltooligosaccharides and starch in *Bacteroides thetaiotaomicron* | Yes (2 m) | 2 |
| Iron complex outermembrane recepter protein | iroN | Essential for iron uptake in bacterial systems | Yes (2 m) | 3 |
| DNA (cytosine-5-)-methyltransferase | DNMT1 | Enables DNA methylation targeting and maintenance during cell division | Yes (Birth) | 4 |
| UTP--glucose-1-phosphate uridylyltransferase | UGP2 | Required to use glucose in anabolic pathways and processes such as glycoprotein folding control, cellular detoxification, and lactation | Yes (2 m) | 5 |
| Enolase | ENO | Receptor for human plasminogen that can promote recruitment of monocytes | Yes (2 m) | 6,7 |
| Transketolase | tktA, tktB | Required to make erythrose-4-phosphate, which is a precursor of aromatic amino acids and vitamins | Yes (6 m) | 8 |
| Glycosyl transferase family 2 | Glycos_transf_2 | Responsible for galactocerebroside synthesis, which is an antigen that triggers Guillain-Barré-Stohl syndrome | Yes (Birth & 2 m) | 9 |
| 6-phosphogluconate dehydrogenase | PGD | Important to produce NADPH and could be an anticancer target | Yes (Birth) | 10,11 |
| Pyruvate formate lyase activating enzyme | pfI | Activates pyruvate-formate lyase | Yes (Birth & 2 m) | 12,13 |
| Formate acetyltransferase | pfID | Catalyzes the nonoxidative generation of formate and acetyl-Coenzyme A | Yes (2 m) | 12,13 |

| | | | | |
|---|---|---|---|---|
| (pyruvate-formate lyase ) | | | | |
| GDPmannose 4,6-dehydratase | gmd | Catalyzes the transformation of GDP-L-mannose to GDP-L-fucose and is required for exopolysaccharide synthesis in *Caulobacter crescentus* | Yes (Birth) | 14,15 |
| Cysteine desulfurase / selenocysteine lyase | sufS | Mobilizes sulfur atoms from cysteine to target proteins during assembly of Fe-S clusters | Yes (6 m) | 16 |
| natB; sodium transport system permease protein | natB | Catalyzes ATP-dependent electrogenic Na+ extrusion in absence of mechanistically coupled proton or K+ uptake | Yes (2 m, 6 m & 24 m) | 17 |
| braS; two-component system, OmpR family, sensor histidine kinase BraS/BceS | braS | Key regulatory element that allows bacitracin and nisin resistance in *Staphylococcus aureus* | Yes (2 m) | 18 |
| ABC-2.P; ABC-2 type transport system permease protein | ABC-2.P | Catalyzes transport of drugs and carbohydrates in bacteria | Yes (2 m) | 19 |
| Putative ABC transport system ATP-binding protein | ABC.CD.A | Associated with higher relative abundances in healthy and adenoma samples when compared to cancer samples | Yes (2 m) | 20 |
| Large subunit ribosomal protein L19 | RPL19 | Upregulation induces endoplasmic reticulum stress and cell death in breast cancer cells | Yes (2 m) | 21 |
| Transketolase, C-terminal domain | Transketolase | Functions as a regulatory molecule binding site | Yes (2 m) | 22 |
| UDP-glucose 6-dehydrogenase | UGDH | Catalyzes the oxidation of UDP-glucose to UDP-glucuronate and could be an anticancer target | Yes (6 m) | 23,24 |

1 Zhang, Y. & Cronan, J. E., Jr. Transcriptional analysis of essential genes of the Escherichia coli fatty acid biosynthesis gene cluster by functional replacement with the analogous Salmonella typhimurium gene cluster. *J Bacteriol* **180**, 3295-3303, doi:10.1128/jb.180.13.3295-3303.1998 (1998).
2 Reeves, A. R., D'Elia, J. N., Frias, J. & Salyers, A. A. A Bacteroides thetaiotaomicron outer membrane protein that is essential for utilization of maltooligosaccharides and starch. *J Bacteriol* **178**, 823-830, doi:10.1128/jb.178.3.823-830.1996 (1996).
3 Clarke, T. E., Tari, L. W. & Vogel, H. J. Structural biology of bacterial iron uptake systems. *Curr Top Med Chem* **1**, 7-30, doi:10.2174/1568026013395623 (2001).
4 Zhang, G. *et al.* Small RNA-mediated DNA (cytosine-5) methyltransferase 1 inhibition leads to aberrant DNA methylation. *Nucleic Acids Res* **43**, 6112-6124, doi:10.1093/nar/gkv518 (2015).
5 Führing, J. I. *et al.* A quaternary mechanism enables the complex biological functions of octameric human UDP-glucose pyrophosphorylase, a key enzyme in cell metabolism. *Sci Rep* **5**, 9618, doi:10.1038/srep09618 (2015).
6 Candela, M. *et al.* Bifidobacterial enolase, a cell surface receptor for human plasminogen involved in the interaction with the host. *Microbiology (Reading)* **155**, 3294-3303, doi:10.1099/mic.0.028795-0 (2009).
7 Wygrecka, M. *et al.* Enolase-1 promotes plasminogen-mediated recruitment of monocytes to the acutely inflamed lung. *Blood* **113**, 5588-5598, doi:10.1182/blood-2008-08-170837 (2009).
8 Harinarayanan, R., Murphy, H. & Cashel, M. Synthetic growth phenotypes of Escherichia coli lacking ppGpp and transketolase A (tktA) are due to ppGpp-mediated transcriptional regulation of tktB. *Mol Microbiol* **69**, 882-894, doi:10.1111/j.1365-2958.2008.06317.x (2008).
9 Gaspari, E., Koehorst, J. J., Frey, J., Martins Dos Santos, V. A. P. & Suarez-Diez, M. Galactocerebroside biosynthesis pathways of Mycoplasma species: an antigen triggering Guillain-Barré-Stohl syndrome. *Microb Biotechnol* **14**, 1201-1211, doi:10.1111/1751-7915.13794 (2021).
10 Hanau, S. & Helliwell, J. R. 6-Phosphogluconate dehydrogenase and its crystal structures. *Acta Crystallogr F Struct Biol Commun* **78**, 96-112, doi:10.1107/s2053230x22001091 (2022).
11 Lin, R. *et al.* 6-Phosphogluconate dehydrogenase links oxidative PPP, lipogenesis and tumour growth by inhibiting LKB1-AMPK signalling. *Nat Cell Biol* **17**, 1484-1496, doi:10.1038/ncb3255 (2015).
12 Stairs, C. W., Roger, A. J. & Hampl, V. Eukaryotic pyruvate formate lyase and its activating enzyme were acquired laterally from a Firmicute. *Mol Biol Evol* **28**, 2087-2099, doi:10.1093/molbev/msr032 (2011).
13 Zelcbuch, L. *et al.* Pyruvate Formate-Lyase Enables Efficient Growth of Escherichia coli on Acetate and Formate. *Biochemistry* **55**, 2423-2426, doi:10.1021/acs.biochem.6b00184 (2016).
14 Jang, M.-H. *et al.* Molecular cloning of the genes for GDP-mannose 4, 6-dehydratase and GDP-l-fucose synthetase from Bacteroides thetaiotaomicron. *Food Science and Biotechnology* **19**, 849-855, doi:10.1007/s10068-010-0120-0 (2010).

15 Herr, K. L. *et al.* Exopolysaccharide production in Caulobacter crescentus: A resource allocation trade-off between protection and proliferation. *PLoS One* **13**, e0190371, doi:10.1371/journal.pone.0190371 (2018).
16 Loiseau, L., Ollagnier-de-Choudens, S., Nachin, L., Fontecave, M. & Barras, F. Biogenesis of Fe-S cluster by the bacterial Suf system: SufS and SufE form a new type of cysteine desulfurase. *J Biol Chem* **278**, 38352-38359, doi:10.1074/jbc.M305953200 (2003).
17 Cheng, J., Guffanti, A. A. & Krulwich, T. A. A two-gene ABC-type transport system that extrudes Na+ in Bacillus subtilis is induced by ethanol or protonophore. *Mol Microbiol* **23**, 1107-1120, doi:10.1046/j.1365-2958.1997.2951656.x (1997).
18 Hiron, A., Falord, M., Valle, J., Débarbouillé, M. & Msadek, T. Bacitracin and nisin resistance in Staphylococcus aureus: a novel pathway involving the BraS/BraR two-component system (SA2417/SA2418) and both the BraD/BraE and VraD/VraE ABC transporters. *Mol Microbiol* **81**, 602-622, doi:10.1111/j.1365-2958.2011.07735.x (2011).
19 Reizer, J., Reizer, A. & Saier, M. H., Jr. A new subfamily of bacterial ABC-type transport systems catalyzing export of drugs and carbohydrates. *Protein Sci* **1**, 1326-1332, doi:10.1002/pro.5560011012 (1992).
20 Ai, D., Pan, H., Li, X., Wu, M. & Xia, L. C. Association network analysis identifies enzymatic components of gut microbiota that significantly differ between colorectal cancer patients and healthy controls. *PeerJ* **7**, e7315, doi:10.7717/peerj.7315 (2019).
21 Hong, M., Kim, H. & Kim, I. Ribosomal protein L19 overexpression activates the unfolded protein response and sensitizes MCF7 breast cancer cells to endoplasmic reticulum stress-induced cell death. *Biochem Biophys Res Commun* **450**, 673-678, doi:10.1016/j.bbrc.2014.06.036 (2014).
22 Lee, H., Deng, M., Sun, F. & Chen, T. An integrated approach to the prediction of domain-domain interactions. *BMC Bioinformatics* **7**, 269, doi:10.1186/1471-2105-7-269 (2006).
23 Arnold, J. M. *et al.* UDP-glucose 6-dehydrogenase regulates hyaluronic acid production and promotes breast cancer progression. *Oncogene* **39**, 3089-3101, doi:10.1038/s41388-019-0885-4 (2020).
24 Zimmer, B. M., Barycki, J. J. & Simpson, M. A. Integration of Sugar Metabolism and Proteoglycan Synthesis by UDP-glucose Dehydrogenase. *J Histochem Cytochem* **69**, 13-23, doi:10.1369/0022155420947500 (2021).
